# Greater than the sum of its parts: combining epigenetic clocks to characterize the association of biological age acceleration and adiposity in young Filipino adults

**DOI:** 10.64898/2026.03.30.26349740

**Authors:** Romaniya S Voloshchuk, Anthony S Zannas, Christopher W Kuzawa, Nanette R Lee, Delia B Carba, Linda S Adair

## Abstract

**Background:** Diverse epigenetic clocks are known to capture health risks associated with increased adiposity, but their estimates have never been combined to represent a holistic estimate of biological age acceleration (BAA). There is also a gap in research using epigenetic clocks to study adiposity in lower-middle income Asian countries.

**Methods and Findings:** Data from 1,745 participants (21.7±0.3 years old, 45% female) of the Cebu (Philippines) Longitudinal Health and Nutrition Survey were analyzed. BAA was calculated using PCHorvath 2, PCHannum, PCPhenoAge, PCGrimAge, PCDNAmTL, and DunedinPACE. After ascertaining suitability for factor analysis (Kaiser-Meyer-Olkin 0.81), factor analysis was used to create PCFactorAge. Analogously, FactorAge was created using Horvath, Hannum, PhenoAge, GrimAge, DNAmTL, and DunedinPACE. BMI, waist circumference (WC), and waist-to-height ratio (WHtR) were used to represent adiposity. Linear regression was used to test the association of each adiposity measure with each BAA measure.

BMI, WC, and WHtR were positively associated with both BAA combinations: 5 kg/m^2^ higher BMI corresponded to 0.097 (p=0.015) standard deviation (SD) increase in FactorAge and 0.099 (p=0.004) SD increase in PCFactorAge; 10 cm increase in WC—with 0.091 (p=0.005) SD increase in FactorAge and 0.094 (p<0.001) SD increase in PCFactorAge; 0.1 increase in WHtR—with 0.164 (p=0.001) SD increase in FactorAge and 0.163 (p<0.001) SD increase in PCFactorAge. Additionally, WHtR was associated with meaningful increases in PhenoAge, PCPhenoAge, PCHorvath 2, PCHannum, PCGrimAge, and DunedinPACE. WC was positively associated with PCHorvath 2, PCHannum, PCPhenoAge, and DunedinPACE. BMI was positively associated with PCHannum, PCPhenoAge, and DunedinPACE.

**Conclusions:** Our study presents a novel approach to creating a BAA estimate using multiple epigenetic clocks and shows that adiposity measures predict this factor in a young Filipino cohort.

## Introduction

Epigenetic clocks use DNA methylation data to estimate biological age—an estimation of age-associated morbidity and mortality risk, which cannot be measured directly. They can help detect elevated morbidity and mortality risk before its clinical manifestation, widening the window of opportunity for healthy lifespan-promoting interventions. This is particularly important in low- and lower-middle-income countries (LMICs), which have high needs for cost-effective preventive healthcare, but remain underrepresented in epigenetic clock research(1). Research from high-and even upper-middle-income countries may not be generalizable to LMICs due to differences in stressors, environmental exposures, and genetics(1–3). The number of epigenetic clocks is growing, and they all capture different but overlapping aspects of biological aging. Using a novel application of factor analysis, we created a robust measure of biological age acceleration using extant well-characterized epigenetic clocks.

Aging is reflected in many molecular mechanisms(4–6), and epigenetic clocks capture variation in related, but distinct, aspects of the aging process, suggesting that a composite measure reflecting their shared variance will present a more complete picture of overall morbidity and mortality risk, and minimize the contributions of biological noise. First-generation epigenetic clocks developed by Horvath(7) and Hannum(8) were trained to predict only chronological age, second-generation clocks, such as PhenoAge,(9) GrimAge,(10) and DNA methylation leukocyte telomere length estimator (DNAmTL)(11) were each trained to predict biological age represented by a different set of aging-related measures of biomarkers, and DunedinPACE(12) was trained on the longitudinal pace of change in aging-related biomarkers (see Table 1). The overlap between genomic sites used by these six clocks is minimal, with any pair of them sharing under 10% of CpGs(13,14), meaning that they estimate biological age from different starting information.

**Table 1:**
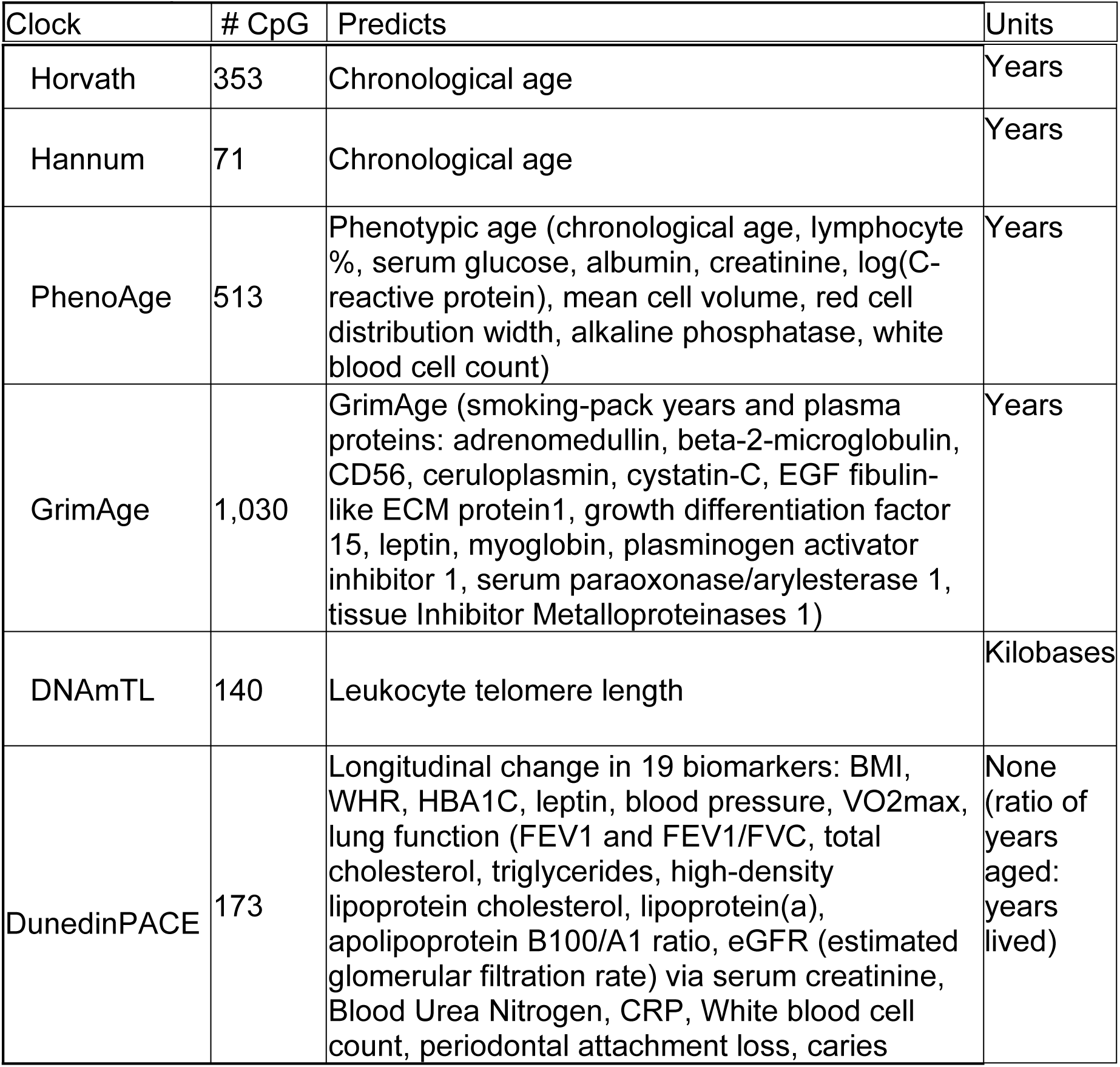
Epigenetic clocks used and their notable features.

| Clock | # CpG | Predicts | Units |
| --- | --- | --- | --- |
| Horvath | 353 | Chronological age | Years |
| Hannum | 71 | Chronological age | Years |
| PhenoAge | 513 | Phenotypic age (chronological age, lymphocyte %, serum glucose, albumin, creatinine, log(C-reactive protein), mean cell volume, red cell distribution width, alkaline phosphatase, white blood cell count) | Years |
| GrimAge | 1,030 | GrimAge (smoking-pack years and plasma proteins: adrenomedullin, beta-2-microglobulin, CD56, ceruloplasmin, cystatin-C, EGF fibulin-like ECM protein1, growth differentiation factor 15, leptin, myoglobin, plasminogen activator inhibitor 1, serum paraoxonase/arylesterase 1, tissue Inhibitor Metalloproteinases 1) | Years |
| DNAmTL | 140 | Leukocyte telomere length | Kilobases |
| DunedinPACE | 173 | Longitudinal change in 19 biomarkers: BMI, WHR, HBA1C, leptin, blood pressure, VO2max, lung function (FEV1 and FEV1/FVC, total cholesterol, triglycerides, high-density lipoprotein cholesterol, lipoprotein(a), apolipoprotein B100/A1 ratio, eGFR (estimated glomerular filtration rate) via serum creatinine, Blood Urea Nitrogen, CRP, White blood cell count, periodontal attachment loss, caries | None (ratio of years aged: years lived) |

It is common to use several epigenetic clocks as outcomes in association studies, which presents several difficulties in interpretation. Firstly, the application of false discovery adjustment is inconsistent across studies, ranging from Bonferroni adjustment(15) to less conservative Benjamini-Hochberg(16) to none(17). Secondly, due to uncertainty about the biological mechanisms driving DNA methylation differences reflected in different epigenetic clocks and differences in their associations with the same exposure(16,18,19), the big-picture interpretation of findings is unclear. Thirdly, on an individual level, the extant epigenetic clocks show a notable margin of error owing to biological noise(20). These challenges can be addressed by combining epigenetic clock estimates using factor analysis(21)—a data reduction method that estimates an unmeasured (latent) variable from measured (indicator) variables using their shared variance, and that can be used to verify that the given data supports this theoretical construct.

While proposing a novel approach to estimating biological age acceleration, this study helps address an inequity in research on adiposity and epigenetic clocks. Studies investigating the relationship between BMI or any other measure of adiposity and epigenetic age in participants from Asian LMICs have yet to be performed. This is an important gap to address, because in populous economically dynamic South and Southeast Asian LMICs, the increase in obesity rates is accelerating(22,23), and projected to continue as the countries grow economically, particularly among the poor(24). In tandem, their years of healthy life lost due to excess adiposity have risen faster than in the rest of the world since 1990(25). These countries are ranked as some of the world’s least prepared to support the growing rates of obesity and obesity-related conditions they are projected to face(26). Studying the relationship between adiposity and widely used epigenetic clocks in young adults can help inform strategies to diminish future disease burden attributable to excess adiposity.

Excess adiposity is a major risk factor for leading contributors to death and disability worldwide(6,27). Positive associations between increased adiposity (especially central adiposity) and aging have been well documented. Aging results in weight gain and fat distribution that favors hormonally-active visceral adiposity, which contributes to metabolic syndrome(28,29). At the same time, individuals with obesity exhibit early signs of aging due to increased systemic inflammation and metabolic dysregulation(29–31). Systemic inflammation and heightened vulnerability to chronic cardiometabolic disease is reflected in white blood cell adaptations through differential DNA methylation(30,32) reflected in epigenetic clocks.

In past work in North American and Western European populations, obesity has been associated with accelerated Horvath, GrimAge, PhenoAge, and DunedinPACE, and telomere shortening(33–37). Body mass index (BMI) and central adiposity measures were shown to have positive linear associations with GrimAge, PhenoAge, and DunedinPACE(18,38,39). Hannum exhibited deceleration following 12 months post-bariatric surgery weight loss(40). In adolescents, high BMI was associated with accelerated GrimAge and DunedinPoAm(19).

While the sensitivity of these epigenetic clocks to adiposity has been widely accepted, most of the evidence supporting this claim is from high-income countries (HICs), collected mostly from people of European and North American descent. The limited evidence from LMICs is mixed. In an age-diverse cohort from Côte d’Ivoire (18-79 yrs), GrimAge acceleration was positively associated with underweight and obesity when compared to study participants whose BMI fell within the normal range, but no such association was detected for Horvath, Hannum, or PhenoAge(41). In a study investigating biological age differences between Ghanaians living in Ghana and Ghanaian migrants living in Western Europe, BMI was negatively associated with biological age in the non-migrant Ghanaians, but no such relationship was observed among the migrants(42). While this could partly be explained by a difference in mean BMI of the two populations, prevalence of other cardiometabolic factors, lifestyle differences, structural differences in the two settings, other unmeasured variables likely contributed to these results.

In this paper, we used factor analysis to estimate biological age acceleration from six epigenetic clocks and tested its association with adiposity in a young Filipino cohort. We used Horvath, Hannum, PhenoAge, GrimAge, DNAmTL, and DunedinPACE because they represent the extant most commonly-used clocks (Table 1). We repeated our analyses using the principal component (PC)-based versions of Horvath, Hannum, GrimAge, PhenoAge, and DNAmTL (PCHorvath 2, PCHannum, PCGrimAge, PCPhenoAge, and PCDNAmTL, respectively), which are increasingly used instead of the original clocks due to improved reliability(20). We then showed the cross-sectional associations of epigenetic clock combinations and the individual clocks used to calculate them with adiposity in a young Filipino cohort.

To our knowledge, factor analysis has not yet been used to combine multiple epigenetic clock estimates of biological age acceleration to create a latent biological age acceleration variable. The approach most similar to ours was taken by Hamlat et al who used structural equation modeling to create a latent variable by combining one epigenetic clock (GrimAge) with measured telomere length and circulating C-reactive protein levels(43). The distinction of our approach is that we identify a latent variable that explains the shared variance across a suite of the most widely used epigenetic clocks, which were individually developed to capture complementary dimensions of the aging process. By identifying the variance shared by these metrics, our factor approach allows for track the biological aging process that is shared across them and may be a more robust representation of biological age acceleration than an individual epigenetic clock.

## Methods

### Population

Data from the 2005 survey round of the Cebu Longitudinal Health and Nutrition Survey (CLHNS) were used. The CLHNS is a community-based birth cohort comprised of index children born between May 1, 1983 and April 30, 1984 in randomly-selected administrative units of Metro Cebu, Philippines, and their mothers(44). We analyzed DNA methylation data from venous blood collection and sociodemograpic data from 1,745 participants (45% female) aged 21.7±0.4 yrs. Relevant questionnaires can be found in https://dataverse.unc.edu/dataset.xhtml?persistentId=hdl:1902.29/11701. Deidentified sociodemographic data can be downloaded from https://dataverse.unc.edu/dataverse/cebu. Other data in this analysis is available at https://github.com/rsvolosh/BioAge_Adiposity.

This study was conducted in accordance with the ethical principles outlined in the Declaration of Helsinki. All procedures involving human subjects were reviewed and approved by the Institutional Review Boards at University of North Carolina at Chapel Hill & University of San Carlos. Written informed consent was obtained from all participants who participated in 2005 – the year the biomarkers used in this study were collected. To obtain data from previous surveys used in this study (household wealth, 1983-2002), verbal consent was obtained by Cebuano-speaking trained staff, because it was sufficient to comply with the regulations in place at that time. This study used de-identified secondary data, accessed January 17, 2025. The authors had no access to information that could identify participants during or after data collection.

Those pregnant at the time of measurement were excluded (n=71) due to challenges interpreting our chosen measures of adiposity in pregnancy, and effects of pregnancy on epigenetic age estimates. Twins (n=21) were excluded because they did not participate in the initial 1983 survey.

### Exposure

Adiposity was measured using BMI, waist circumference (WC), and waist-to-height-ratio (WHtR). After initial analyses, measures were standardized using sex-specific within-sample z-scores to facilitate comparing coefficients. Z-scores were calculated for males and females separately for the following reasons: (1) in our sample, the distributions (means and standard deviations) were different for males and females, as shown in Table 2, and (2) healthy ranges for WC differed for females and males (80 cm and 90 cm for WC, per the International Diabetes Federation cut-off for South and East Asians(45), respectively).

**Table 2:**
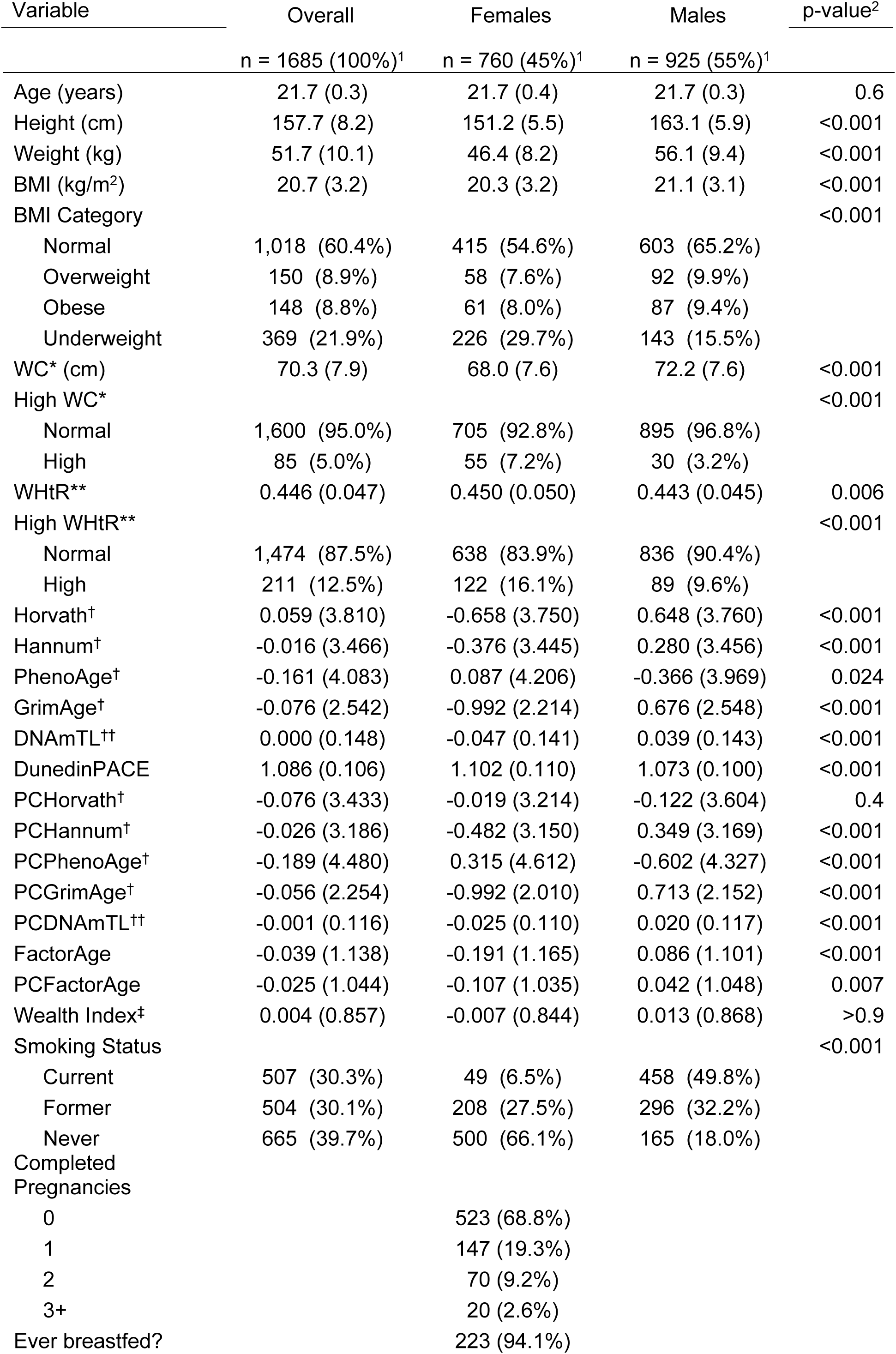
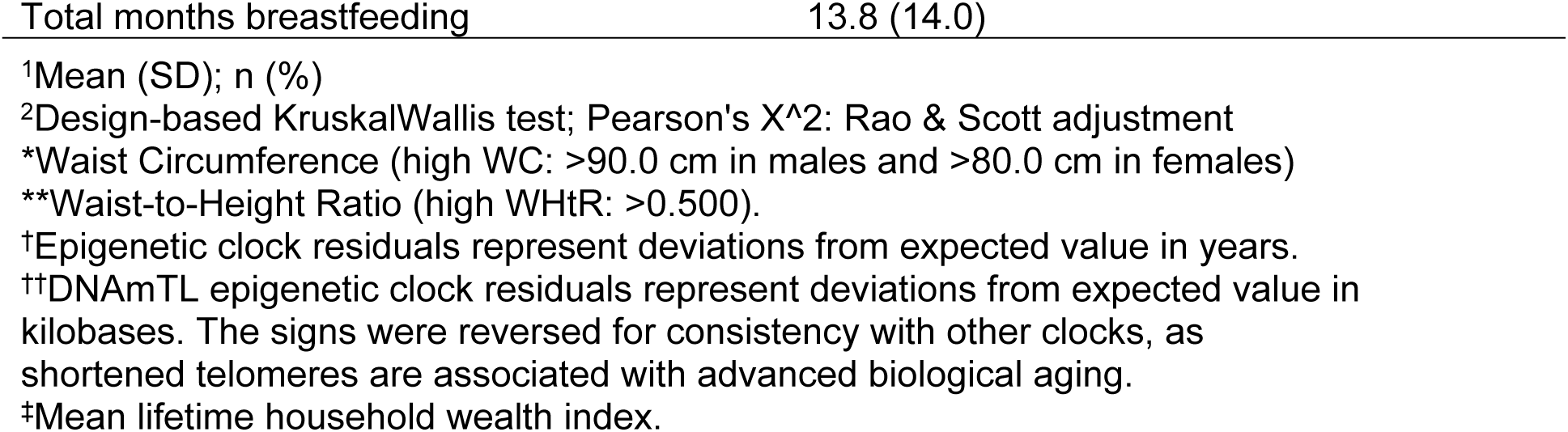
Sample Characteristics. Residuals were used for epigenetic clocks.

Waist measurements missing in the absence of pregnancy (n=2) were imputed using multiple imputation by chained equations (MICE), using the “mice” package in R(46).

### Outcome

#### DNA methylation & epigenetic clock estimates

Venous blood samples from overnight-fasted participants were collected in the morning during a home visit by trained CLHNS data collectors. Genomic DNA was (1) extracted from dried blood spots using standard protocol, (2) concentrated using a Vacufuge Plus vacuum concentrator (Eppendorf), (3) treated with sodium bisulphite to determine which CpG sites were methylated (Zyme EZDNA, Zymo Research, Irvine, CA), and (4) applied to the Illumina Infinium MethylationEPIC BeadChip under standard conditions (Illumina Inc., San Diego, CA). Background subtraction and colour correction were performed using Illumina Genome Studio with default parameters. To minimize batch effects, samples were randomized on runs by baseline neighbourhood; samples were randomly assigned to plate, chip, and row. Quality control involved (1) confirming participant sex and replicate status, and (2) quantile normalization using lumi on all probes, including SNP-associated and XY multiple binding probes(47). Technicians were masked from information about participant characteristics. To maximize the number of sites available for the epigenetic age calculator, probes with detection p values above 0.01 were called NA for poor performing samples only but were otherwise retained(48). A subset of 30,084 probes from the EPIC array (including rows with Illumina IDs only if not found on the EPIC array) was then constructed and uploaded with accompanying age, sex, and tissue information to the online epigenetic clock calculator (http://labs.genetics.ucla.edu/horvath/dnamage/) using the ‘Advanced analysis’ option to calculate epigenetic age using Horvath, Hannum, PhenoAge, GrimAge, and DNAmTL clocks. Additionally, principal component-based versions of these clocks were used to calculate epigenetic age, following methods and materials described by Higgins-Chen et al.(20) The Belsky PACE clock was calculated using the DunedinPACE R package (https://github.com/danbelsky/DunedinPACE). Peripheral blood cell composition was imputed from the DNAm data using previously described methods(49,50). Except for DunedinPACE, clock values reported in Table 2 were residualized on chronological age to obtain measures of biological age acceleration uncorrelated with age. DunedinPACE was instead trained to predict the change in 19 markers of organ system integrity and represents estimated rate of aging [(biological years aged)/(chronological year lived)], thus necessitating no residualization on chronological age. GrimAge and PCGrimAge were missing and imputed using MICE(51) for one participant.

#### Epigenetic Clock Combinations

In preparation for factor analysis, residuals for all clocks were standardized, adjusting for chronological age for all but DunedinPACE(12). Then, factor analysis was performed twice—once on the original clocks (Horvath, Hannum, GrimAge, PhenoAge, DNAmTL, DunedinPACE), and once on their corrected versions per Higgins-Chen et al (PCHorvath 2, PCHannum, PCGrimAge, PCPhenoAge, PCDNAmTL, DunedinPACE). Note: the motivation for creating PC-based (corrected for biological noise) epigenetic clock versions was to bolster their reliability; DunedinPACE had high reliability in its non-PC form (comparable to PC-based clocks)(12,20), so we treated DunedinPACE as its own PC-based version.

#### Factor Analysis

Exploratory factor analysis (FA)(21) was conducted to create epigenetic clock combinations using the “psych” package in R(52). The suitability of data for FA was assessed using the Kaiser, Meyer, Olkin (KMO) test(53). Then, eigenvalues and scree plots were used to test for the optimal number of factors for the clocks and their PC-based versions. In the non-PC factor analysis, Horvath, Hannum, PhenoAge, GrimAge, DNAmTL, and DunedinPACE were included. In the PC factor analysis, PCHorvath 2, PCHannum, PCPhenoAge, PCGrimAge, PCDNAmTL, and DunedinPACE were included. Bartlett method was used to generate factor scores.

### Covariates

Analyses were adjusted for biological sex (sex was defined as sex assigned at birth; no information about change in gender identity was available), underweight status, blood cell type proportions, average lifetime household wealth, smoking, and for females, the number of completed pregnancies, and total months spent breastfeeding. Models including WC were additionally adjusted for height.

Since underweight (BMI < 18.5 kg/m^2^) is associated with adverse health outcomes, in contrast to people whose BMI was above 18.5 kg/m^2^, we did not expect higher adiposity to be associated with higher biological age among people who were underweight. Because 22% of CLHNS participants were underweight at the time of data collection, analyses were adjusted by interacting the adiposity variable with a binary underweight variable. This approach is analogous to using linear splines with a knot at 18.5. In models where this interaction was not statistically significant (alpha<0.05) and did not improve model fit (ANOVA, alpha<0.05), the interaction term was removed, but underweight status was kept as a covariate.

Cell type proportions were calculated per Houseman et al(49), and adjusted for to ensure that differences in methylation observed were not due to differences in blood cell composition.

Household socioeconomic status (concurrent and during development) is an important determinant of weight status(54) and health outcomes independent of weight status(15,55,56). Mean lifetime household wealth index was used as proxy for socioeconomic status. Household wealth index at 1983 (birth – 0 yrs), 1991 (8 yrs), 1994 (11 yrs), 1998 (15 yrs), 2002 (18 yrs), and 2005 (21 yrs) surveys was calculated using the first principal component extracted from a polychoric principal component analysis performed on the possession of household assets (see S1 Table for details), harmonized, and standardized to reflect cross-sectional differences in household wealth relative to the other participants in the study (57,58). Then, missing values were imputed using MICE (n=64 missing at one survey; n=8 missing at two surveys). Then, the factor scores were averaged for each participant to yield a variable representing average lifetime household wealth.

Smoking status is a determinant of adiposity(59,60) and biological age independent of adiposity(15,34,61). A three-level categorical variable (never vs. former vs. current smoker) was used.

Parity is associated with adiposity(62) and biological age acceleration(17). Breastfeeding is associated with post-partum weight change(63), and has been associated with some positive maternal health outcomes(64–67), which could be reflected in biological age measures. Hence, we adjusted the models by number of completed pregnancies interacted with total months spent breastfeeding. 237 females were parous, of which 90 had more than one completed pregnancy and 223 breastfed at least one child. Miscarriages (n=11) were not counted as completed pregnancies, but stillbirths (n=5) were. Twin pregnancies (n=3) were counted as one pregnancy. Breastfeeding months were calculated from the total time breastfeeding from all completed pregnancies.

WC, unlike BMI or WHtR, does not account for height but is partially determined by it(68). Adult height could also represent exposure to early-life stressors, which are associated with biological age(56), making it a potential confounder in models using WC as the exposure. We added height interacted with sex to the fully adjusted WC models.

### Modeling Approach

Multivariate linear regression was used to separately test the association of each adiposity measure with each epigenetic measure of age and each of their combinations as described above. Model 1 was adjusted for only sex interacted with adiposity measure (females were the referent group). In Model 2, all other covariates were added, including the interaction between underweight status and adiposity. If the interaction terms which included adiposity were statistically significant (p<0.05), Model 2 was the final (fully adjusted) model. If these interactions were not statistically significant, they were removed one-by-one, difference in model fit was assessed using ANOVA, and if the interaction terms did not significantly improve model fit (p<0.05), the reduced model was chosen to be the fully adjusted model; otherwise, additional sex-stratified models were run, testing the need for the interaction term between adiposity and underweight using the aforementioned method. For models with statistically significant underweight-adiposity interactions, predictive margins were created using the “predict” function in R and plotted. Benjamini-Hochberg correction for multiple testing was performed for the individual epigenetic clocks (PCHorvath, PCHannum, PCPhenoAge, PCGrimAge, PCDNAmTL, and DunedinPACE)(69). Analogous adjustment was performed for the original versions of these clocks. Goodness of fit was calculated using “modelsummary” function in R(70). Regression coefficients of interest were compared using methods outlined in Clogg, Petkova, and Haritou(71). G*Power(72) was used to complete sensitivity power analysis for multivariate regression with alpha = 0.05, 0.80 power, and 14 degrees of freedom; *η*^2^ was reported in addition to f^2^ as suggested by Correll et al(73). Code used in this article is available at https://github.com/rsvolosh/BioAge_Adiposity. Power analysis showed that a small effect size could be detected using our data.

## Results

### Sample characteristics

Most participants were in the “normal” BMI category (18.5kg/m^2^ ≤ BMI < 23kg/m^2^) as defined by cut-points recommended for Asia-Pacific by the World Health Organisation(74–76), with overweight and obesity being more common in males than females, and underweight being more common in females than in males. Females were more likely to have a high WC and WHtR than males. The higher proportion of females than males having a high WC was not fully explained by parity (S4 Table), with nulliparous females still having higher rates of high WC than males (5.5% vs. 3.2%). Smoking rates markedly differed by sex. 523 females were nulliparous, 237 had given at least one birth by the time of the study,; 94% of parous females had breastfed, with a mean breastfeeding duration of 13.4 months (Table 2).

### Correlations of residuals and suitability for factor analysis

Characteristics of epigenetic clocks used here are described in Table 1. Their age estimates were mostly highly correlated (Table 3). Original, non-PC-based versions showed significantly smaller correlations than their PC versions (Table 3), ranging from 0.003 (DunedinPACE and Horvath) to 0.481 (Hannum and DNAmTL). Correlations among epigenetic clock residuals were similar for males and females and ranged from 0.003 (DunedinPACE vs. Horvath) to 0.843 (PC Horvath 2 vs. PC Hannum). PC clocks were more highly correlated with each other than non-PC clocks (min: 0.003 vs. 0.269; median: 0.543 vs. 0.277; max: 0.843 vs. 0.481). Epigenetic clocks showed moderately high correlations with their PC versions (0.622-0.811), except Horvath vs. PC Horvath 2 (0.382) (Table 3).

**Table 3:**
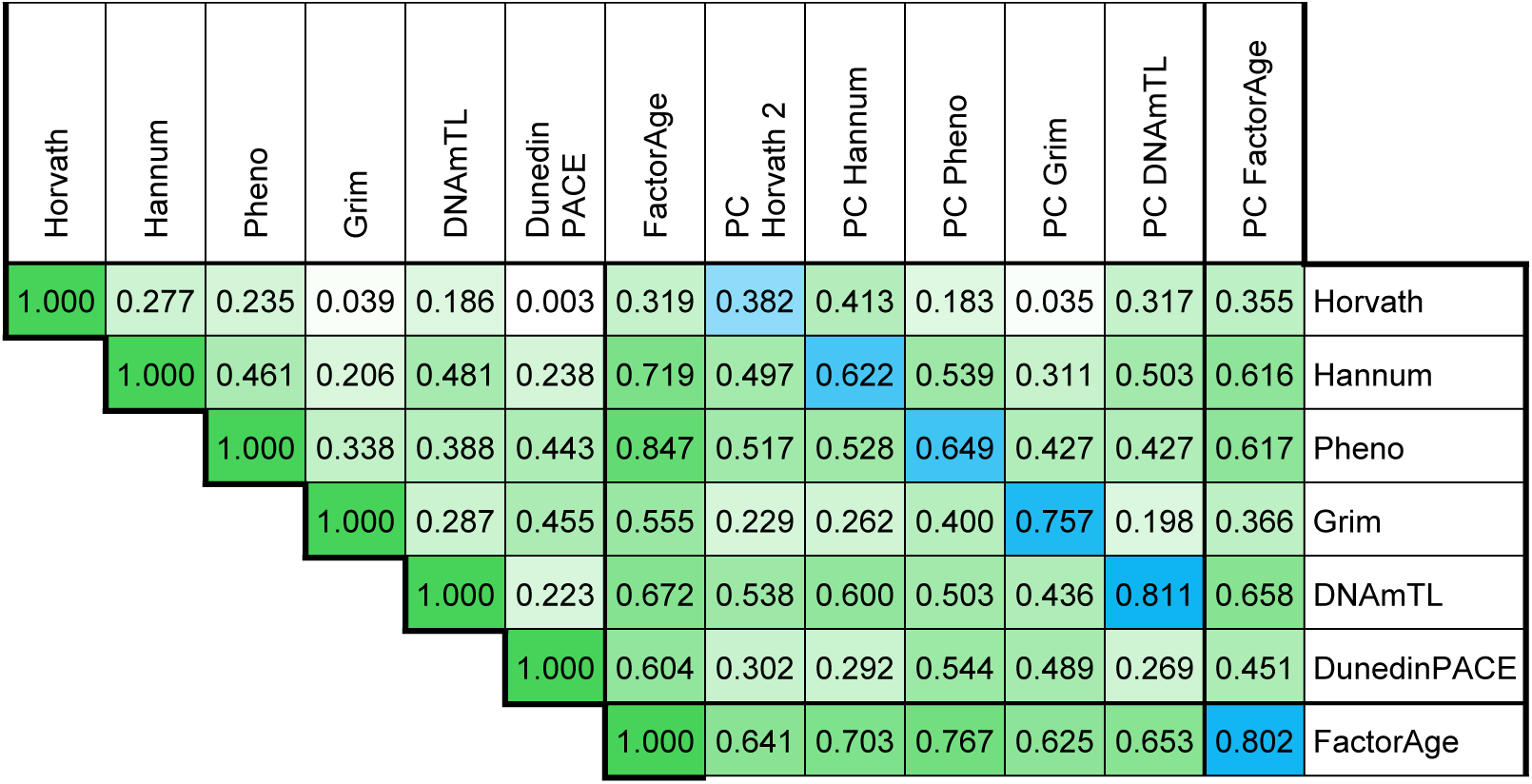
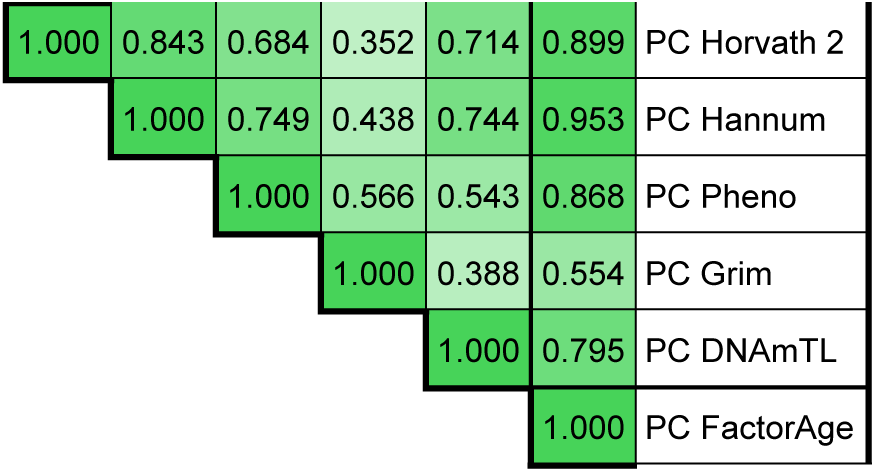
Correlations of epigenetic clocks residualised on chronological age.

| Horvath | Hannum | Pheno | Grim | DNAmTL | Dunedin PACE | FactorAge | PC Horvath 2 | PC Hannum | PC Pheno | PC Grim | PC DNAmTL | PC FactorAge |  |
| --- | --- | --- | --- | --- | --- | --- | --- | --- | --- | --- | --- | --- | --- |
| 1.000 | 0.277 | 0.235 | 0.039 | 0.186 | 0.003 | 0.319 | 0.382 | 0.413 | 0.183 | 0.035 | 0.317 | 0.355 | Horvath |
|  | 1.000 | 0.461 | 0.206 | 0.481 | 0.238 | 0.719 | 0.497 | 0.622 | 0.539 | 0.311 | 0.503 | 0.616 | Hannum |
|  |  | 1.000 | 0.338 | 0.388 | 0.443 | 0.847 | 0.517 | 0.528 | 0.649 | 0.427 | 0.427 | 0.617 | Pheno |
|  |  |  | 1.000 | 0.287 | 0.455 | 0.555 | 0.229 | 0.262 | 0.400 | 0.757 | 0.198 | 0.366 | Grim |
|  |  |  |  | 1.000 | 0.223 | 0.672 | 0.538 | 0.600 | 0.503 | 0.436 | 0.811 | 0.658 | DNAmTL |
|  |  |  |  |  | 1.000 | 0.604 | 0.302 | 0.292 | 0.544 | 0.489 | 0.269 | 0.451 | DunedinPACE |
|  |  |  |  |  |  | 1.000 | 0.641 | 0.703 | 0.767 | 0.625 | 0.653 | 0.802 | FactorAge |

|  |  |  |  |  |  |  |
| --- | --- | --- | --- | --- | --- | --- |
| 1.000 | 0.843 | 0.684 | 0.352 | 0.714 | 0.899 | PC Horvath 2 |
|  | 1.000 | 0.749 | 0.438 | 0.744 | 0.953 | PC Hannum |
|  |  | 1.000 | 0.566 | 0.543 | 0.868 | PC Pheno |
|  |  |  | 1.000 | 0.388 | 0.554 | PC Grim |
|  |  |  |  | 1.000 | 0.795 | PC DNAmTL |
|  |  |  |  |  | 1.000 | PC FactorAge |

KMO Measure of Sampling Adequacy (MSA) showed that all clock values were between 0.68 and 0.87. Overall MSA was 0.73 for non-PC and 0.81 for PC clocks (see S2 Table). Scree plots showed only one factor with an eigenvalue greater than 1 (see S1 Fig).

### Factor analysis

Factor score adequacy measures indicated sufficiency for the single-factor solutions in PC and non-PC scores (see S1 Fig and S3 Table). In non-PC clocks, the factor score was driven by PhenoAge (factor loading 0.74) and Hannum (factor loading 0.63). For the PC versions, all clocks except for DunedinPACE and GrimAge loaded at 0.78 or higher.

### Linear Model Results

Our sample size was adequate to detect the effect sizes reported in the studies cited in the introduction: the critical F for multiple linear regression was 1.70 and detectable effect size was small (f^2^=0.01, or *η*^2^=0.01); for sex-stratified analyses, critical F = 1.73 for females and 1.84 for males with f^2^ = 0.02, or *η*^2^=0.02 for both.

The only outcome for which exposure interaction with sex or underweight status were statistically significant or significantly improved model fit was DunedinPACE (interactions with underweight were omitted from Table 5A to improve readability). Goodness of fit statistics for Models 1, 2, and final can be found in S1 File.

DunedinPACE was the only clock showing sex-specific associations with adiposity. In sex-stratified models, it showed a strong positive relationship with all adiposity measures in females, but this relationship was weaker in males (Table 5B). DunedinPACE also showed a differential relationship with adiposity by underweight status (see Table 5B and predictive margins in Fig 1). For WHtR, in unadjusted models, in addition to DunedinPACE the GrimAge coefficient was significant with FDR adjustment, while Hannum and DNAmTL coefficients were statistically significant without FDR adjustment (S1 File). No statistically significant associations were found for Horvath.

**Fig 1:**
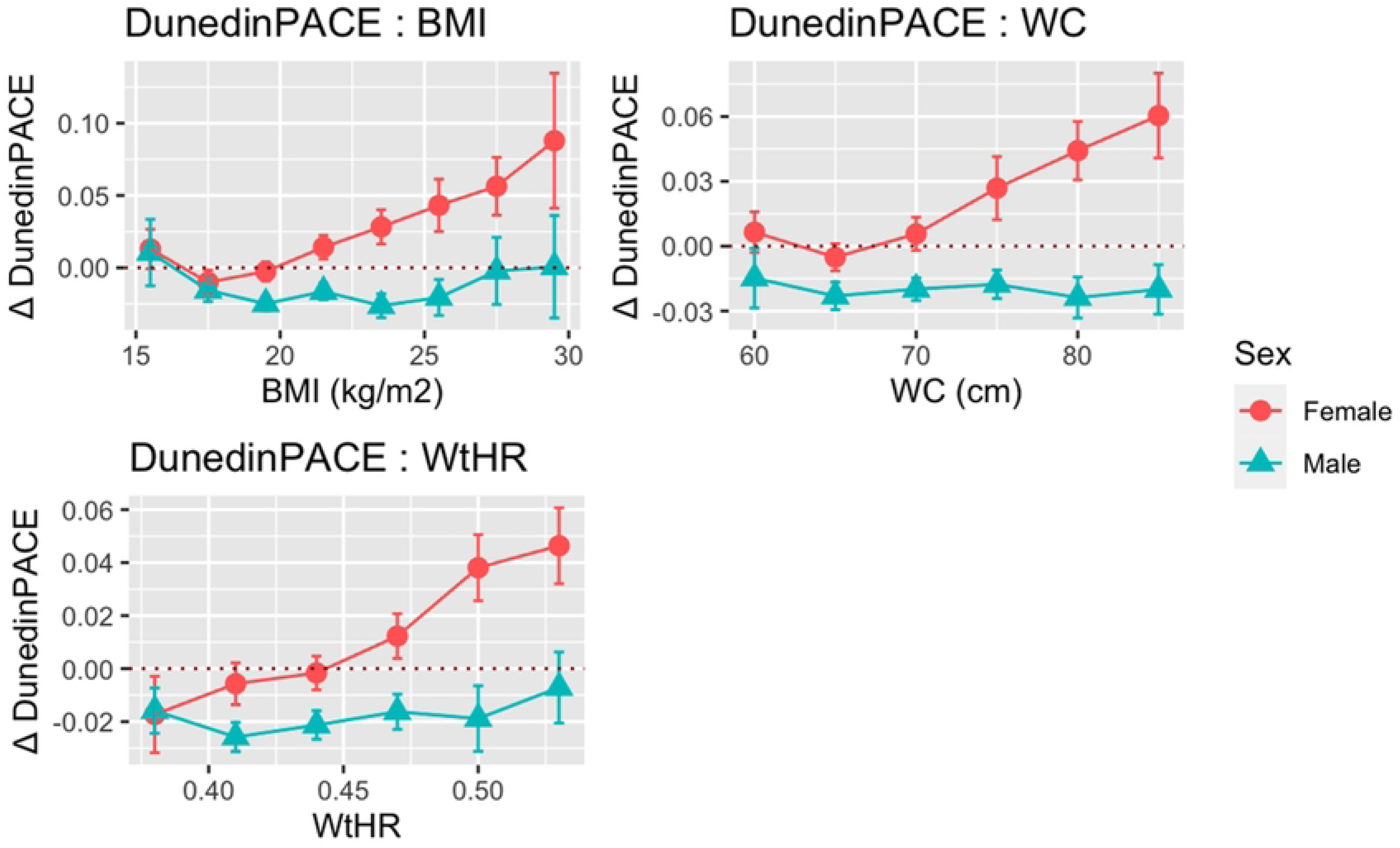
Predictive margins for DunedinPACE. Regression coefficients presented for increase of 1SD BMI, WC, and WHtR; outcomes presented in changes of standard deviation.

With the exception of PCDNAmTL, which did not show any statistically significant correlations with any adiposity measures, PC versions of epigenetic clocks showed stronger associations with BMI, WC, and WHtR (Table 5): PCPhenoAge and PCHannum showed associations with all three measures, PCHorvath 2 – with WC and WHtR, and PCGrimAge with WHtR only.

FactorAge and PCFactorAge were associated with all adiposity measures at a statistically significant level, showing similar regression coefficients and statistical significance levels, despite FactorAge being comprised of estimates most of which showed lower associations with adiposity, and despite having different factor loadings for analogous clocks (Table 6).

Both combinations were strongly associated with BMI, WC, and WHtR (p < 0.01). Their coefficients for BMI and WC were nearly identical (Table 6); the coefficient for WHtR appeared to be higher for FactorAge than PCFactorAge (0.164, p=0.001 vs. 0.140, p=0.003), but when empirically compared,(71) the difference between these coefficients was not statistically significant (d = −0.023, standard error = 0.064).

Goodness of fit for unadjusted and adjusted models can be found in (S1 File). Briefly, for Factor and PC Factor models, adjusted R^2^ was above 0.4, and above 0.2 for all clocks except for Horvath (R^2^ = 0.1). Models with PC clocks as outcomes had better goodness of fit – R^2^ was above 0.4, except for PCHorvath 2 and PCHannum, for which it was still above 0.3 for all adiposity measures.

When comparing statistically significant regression coefficients to find if they differed for models using Factor, PCFactor, or individual epigenetic clocks as outcome, we found that the differences in these coefficients was not statistically significant, with the exception of DunedinPACE, whose estimates for non-underweight females were consistently higher than those of Factor or PCFactor, except for non-underweight males (S5 Table).

## Discussion

In this study we demonstrated the feasibility of calculating a holistic measure of biological age acceleration using factor analysis (FactorAge and PC FactorAge) which treats biological age as a latent variable. In light of the pressing need to better understand the relationship between adiposity and biological age in the populous Asian LMICs, we tested how sensitive FactorAge was to three adiposity measures, and how it compared to the individual epigenetic clocks used to estimate FactorAge in a young Filipino birth cohort. We found higher adiposity to be similarly associated with biological age acceleration estimated using Factor and PC Factor; most individual PC clocks showed stronger associations than non-PC; only DunedinPACE captured sex-specific non-linear associations.

Biological age, which refers to the decline in functional capacity and increased risk of death and disease, is inherently difficult to measure(9,48). Epigenetic clocks can be thought of as indicators of biological age, reflecting epigenetic adaptations influenced by functional decline and chronic disease risk. Factor analysis is a method designed to capture variables which cannot be measured directly, and has been successfully applied in biological contexts, facilitating the study of developmental origins of health and disease(77). One of its advantages is its facilitation of verifying that the data supports the hypothesis that it is appropriate to combine estimates of several epigenetic clocks, meaning that they are, in fact, indicators of one latent variable.

Factor analysis results supported combining epigenetic clocks to create Factor and PC Factor: MSA for the six clocks chosen for this study in their original form was 0.73, and 0.81 for their PC versions. KMO test results showing MSA higher than 0.50 is considered acceptable, with most real-world cases expected fall between 0.70 and 0.90(53).

The latent variable created using factor analysis captures the variance shared by the different epigenetic clocks (communality shows the proportion of variance of each measure explained by the latent variable). The remaining variance of each epigenetic clock estimate is partly due to technical error, including biological noise, and variability in the proxy for biological age used to train the clock that is not shared by proxies used to create other epigenetic clocks (e.g. variability in telomere length uncorrelated with chronological age). Higher communalities in factor analysis using measures more robust to biological noise (PC-based clocks) shown in Table 4 support the claim that there is a portion of variance in epigenetic clock estimates of biological age acceleration that is due to technical error, and this error can be mitigated using factor analysis. Factor analysis helps leverage the strength of each epigenetic clock and reinforces their correlations.

**Table 4:** Factor loadings for single factor solution.

A: Original (non-PC) Factor Analysis
|  | Loading | Communality |
| --- | --- | --- |
| Horvath | 0.28 | 0.08 |
| Hannum | 0.63 | 0.40 |
| PhenoAge | 0.74 | 0.54 |
| GrimAge | 0.48 | 0.23 |
| DNAmTL | 0.58 | 0.34 |
| DunedinPACE | 0.53 | 0.28 |

B: PC Clock Factor Analysis
|  | Loading | Communality |
| --- | --- | --- |
| PCHorvath 2 | 0.89 | 0.78 |
| PCHannum | 0.94 | 0.89 |
| PCPhenoAge | 0.79 | 0.63 |
| PCGrimAge | 0.49 | 0.24 |
| PCDNAmTL | 0.78 | 0.61 |
| DunedinPACE | 0.38 | 0.14 |

Loading refers to factor loadings extracted in this solution.
Communality refers to the portion of the variable explained by the factor.

**Table 5:**
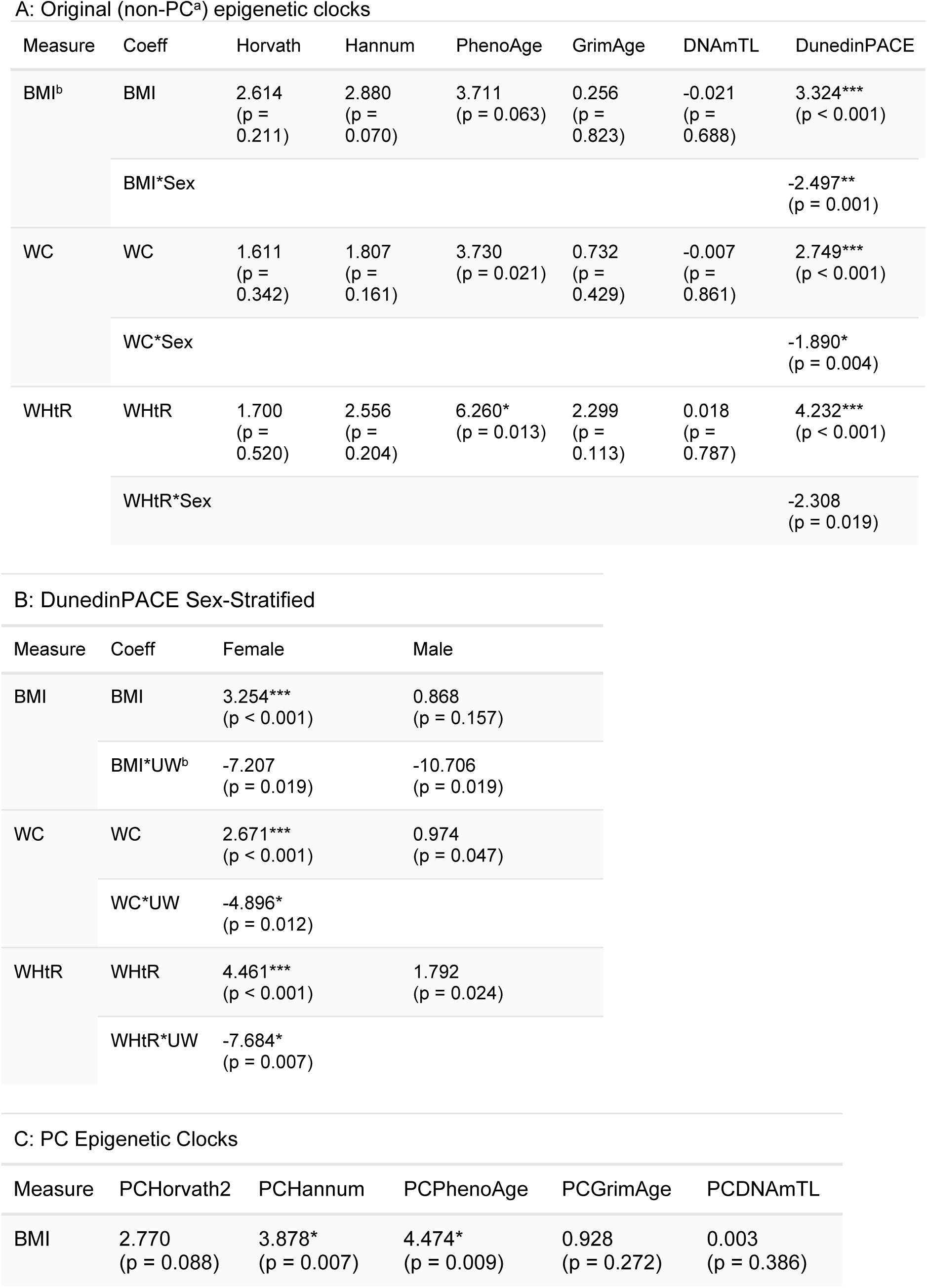
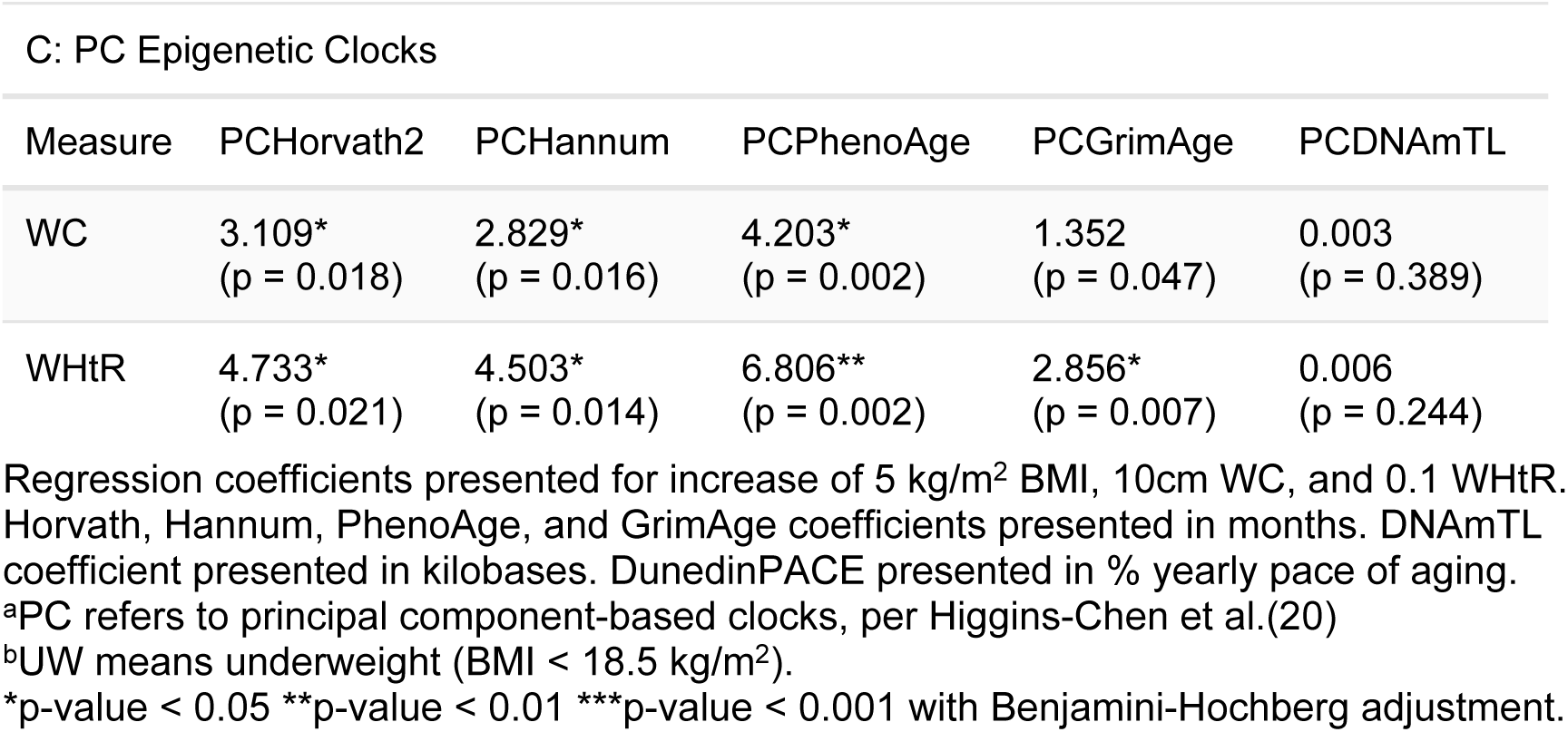
Linear regression results from individual epigenetic clocks.

**Table 6:** Linear regression results for combinations of clocks.

| Measure | FactorAge | PCFactorAge |
| --- | --- | --- |
| BMI | 0.097*<br>(p = 0.015) | 0.097**<br>(p = 0.009) |
| WC | 0.091**<br>(p = 0.005) | 0.085**<br>(p = 0.004) |
| WHtR | 0.164**<br>(p = 0.001) | 0.140**<br>(p = 0.003) |

Regression coefficients presented for increase of 5 kg/m<sup>2</sup> BMI, 10cm WC, and 0.1 WHtR; outcomes presented in changes of standard deviation.
\*p-value < 0.05 \*\*p-value < 0.01 \*\*\*p-value < 0.001

Though most of our cohort did not have overweight or obesity at age 21, people tend to gain weight as they age, and it is important to identify those at risk of developing metabolic syndrome early to increase the window of opportunity for preventive interventions. Most of our cohort (61%) was stunted at 24 months(78), indicating high levels of early-life undernutrition. Undernourishment in early life may increase their risk of obesity, hypertension, and heart disease – growing health issues in the Philippines and LMICs at large(79–82). In LMICs with dynamic economies, as people’s lifestyles become more sedentary and their consumption of calorie-dense processed and ultra-processed foods increases, so do their obesity rates(83–85). Epigenetic clocks facilitate early detection of morbidity risk brought on by such environmental exposures.

Though the measurements used in this study were taken when the study participants were young (21.7±0.3 yrs) and relatively lean, with low rates of overweight and obesity (Table 2), we found statistically significant associations between adiposity and biological age. FactorAge and PCFactorAge showed positive correlations with BMI, WC, and WHtR. For FactorAge, this relationship appears to have been driven by Hannum, PhenoAge, and DunedinPACE, as both their factor loadings were comparatively high (Table 4A), and they were individually correlated with adiposity measures. In line with the expectation that PC versions of epigenetic clocks estimate biological age more precisely than their original counterparts due to reductions in biological noise, factor loadings for all PC clocks were higher than their original counterparts, indicating that the latent variable was explaining a higher portion of variability in PC-based estimates.

We found that, overall, PC versions of epigenetic clocks were all more sensitive to adiposity. This lines up with our expectation, given that they are robust to biological noise because they leveraged the fact that methylation levels across different CpGs co-vary and using their principal components instead of single CpGs. With the exception of PCDNAmTL, all PC-based clocks were associated with at least one measure of adiposity. In the paper describing DNAmTL, the authors showed that the DNAmTL was less strongly associated with adiposity (BMI, WHR) than with ethnicity, education, or smoking; they also remarked that DNAmTL was often inferior to other epigenetic clocks in predicting many important age-related traits.(11) Considering that the cohort in this study was on the extreme low of both age and BMI represented in its the training data, the fact that DNAmTL and PCDNAmTL did not show any associations with adiposity is not surprising.

Comparing regression coefficients for models using Factor, PCFactor, and individual epigenetic clocks as outcomes showed that, overall, their coefficients were similar (S5 Table), which supports the hypothesis that they are all telling the same story with regards to adiposity: generally, increased adiposity is associated with accelerated biological aging, and combining clocks does not yield unexpected results. DNAmTL and PCDNAmTL models had regression coefficients closer to the null, which was expected given the lack of associations we found between adiposity and DNA methylation estimators of telomere length. Due to sex-specific non-linear associations unique to DunedinPACE, it was impossible to empirically compare its estimates, for the whole cohort, but among the non-underweight females the association of every adiposity measure and DunedinPACE was higher than with Factor or PCFactor—an expected result, given that DunedinPACE showed some of the lowest correlations with other epigenetic clocks, represents the pace of biological aging, not a proxy for biological age. However, because the drivers of faster pace biological aging overlap with the drivers of biological age in relation to chronological age (the difference between predicted and actual age), including it in the estimation of a latent biological age acceleration variable was warranted.

When comparing standardized beta coefficients by adiposity measure, we did not find them statistically significantly different for any clock (S6 Table). For WC and WHtR, we also found PC-based clocks to estimate very similar age acceleration measures associated with a one standard deviation increase in the metric (see Tables 7, 8, S6 Table). Overall, in addition to the combinations of clocks, PCHannum, PCPhenoAge, and DunedinPACE showed the highest statistically significant correlations with BMI, WC, and WHtR in our cohort. PhenoAge was trained to predict a score based on clinical blood markers associated with various aspects of aging, many of which are altered in a state of excess adiposity, such as C-reactive protein glucose concentrations; the training of DunedinPACE included even more such biomarkers (including blood pressure, total cholesterol, leptin, WHR, BMI, CRP). Hannum, however, is a first-generation clock that was trained only to predict chronological age; yet it shows stronger associations with adiposity than GrimAge – a second-generation clock, which uses over 1,000 CpGs and was trained to predict blood proteins and smoking-pack years.

**Table 7:**
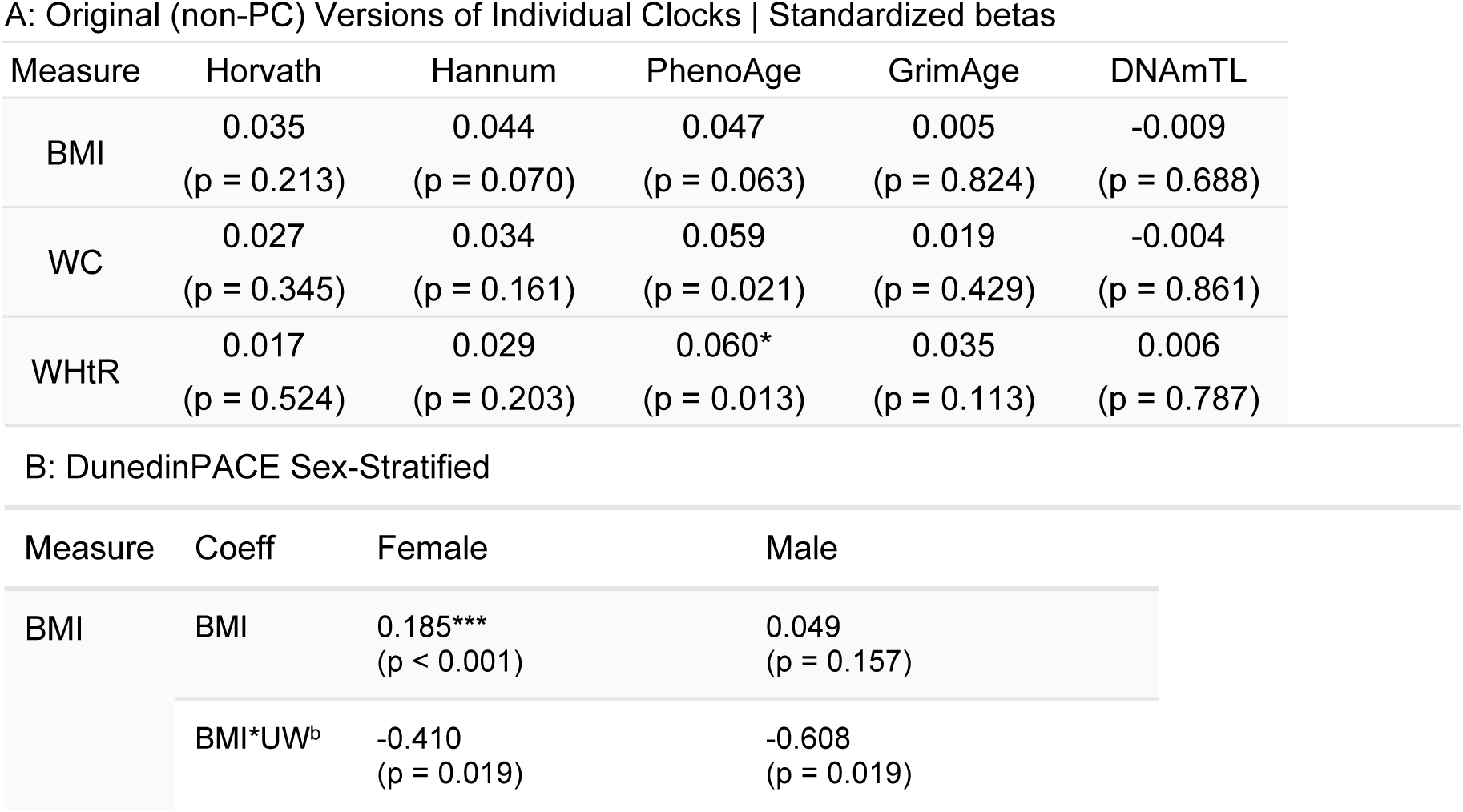
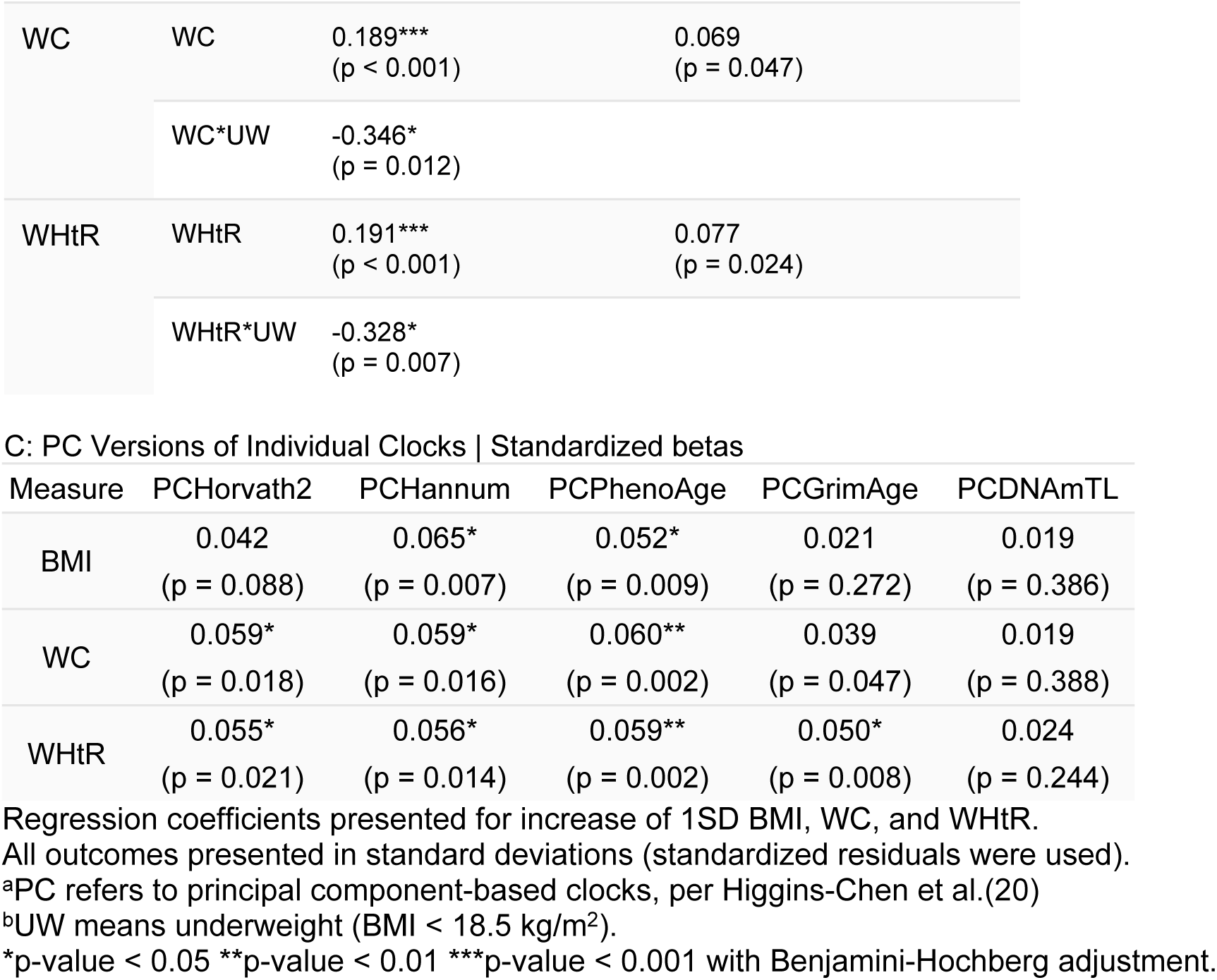
Standardized regression coefficients for adiposity measure z-scores for individual epigenetic clocks.

**Table 8:** Standardized betas for adiposity measure z-scores for combination epigenetic clocks.

| Measure | FactorAge | PCFactorAge |
| --- | --- | --- |
| BMI | 0.062*<br>(p = 0.015) | 0.061**<br>(p = 0.009) |
| WC | 0.072*<br>(p = 0.005) | 0.067**<br>(p = 0.004) |
| WHtR | 0.078**<br>(p = 0.001) | 0.067**<br>(p = 0.003) |

Regression coefficients presented for increase of 1SD BMI, WC, and WHtR;
outcomes presented in changes of standard deviation.
\*p-value < 0.05 \*\*p-value < 0.01 \*\*\*p-value < 0.001

Consistent with the literature, males appeared biologically older than females according to most clocks (Table 2), with FactorAge and PCFactorAge following the same pattern(86,87). It is important to note that males were much more likely to smoke, with half of the males being current smokers, compared with 6.5% of females. However, smoking alone did not explain this sex difference.

DunedinPACE showed sex-specific associations of adiposity and pace of aging. It was highly sensitive to all adiposity measures in females. It is notable that these coefficients are similar, and, when compared empirically, do not differ at a statistically significant level (S6 Table). In males, only WHtR was associated with increased DunedinPACE; its association with WC was statistically significant only without FDR. Higher level of physical fitness in the males could help explain the lack of association between BMI and DunedinPACE: higher BMI could mean higher level of fat or musculature; DunedinPACE was trained to predict the change in 19 markers of organ system integrity, which included grip strength and cardiovascular fitness, measured by VO_2_Max, and could thus capture the anti-aging benefits of adequate physical activity(88).

Though females were more likely than males to be underweight, they were also more likely to have a high WC, even if they were nulliparous. One possible explanation could be that males were likelier to be physically active: they were likelier to have physically demanding jobs (e.g. construction) and to play sports(89–91). Contrarily, thinness was socially desirable for females(92). With relatively sedentary lifestyles, the body fat percentage could have been higher than expected given their BMI in the females, but not males.

This study has several limitations. Though we used the six most popular epigenetic clocks in similar research, there are other epigenetic clocks we did not test that may offer new insights. We also present only one approach to factor analysis, but there may be other reasonable approaches to combining clocks (for example, removing epigenetic clock residuals with low loadings or testing different epigenetic clock combinations). It is difficult to clearly explain what the association coefficients mean in combination clocks without using the cohort as a reference, necessitating extra steps to effectively communicate the findings to the public. Our study population included only young Filipinos, and more research is needed in ethnically and age-diverse populations, especially ones underrepresented in the data used to train epigenetic clocks(1–3). By their nature, none of our adiposity measures give exact estimates of the amount of total, ectopic or visceral fat, musculature, or body water. Lastly, we did not control for months postpartum because, due to their young age and high rates of breastfeeding, breastfeeding duration was a sufficiently good substitute. By nature of our analyses, we could not establish causality or elucidate any biological mechanisms implicated in our results.

The strength of this study lies in its comparability with existing and future research due to the variety of common cost-effective measures of adiposity and several epigenetic clocks, which adequately represent the extant first- and second-generation clocks widely used in biological aging studies. Another strength is that we repeated our analyses with the newer PC epigenetic clocks. Presenting the results from both maximizes the comparability of our results with existing and future research. We offer evidence that combining epigenetic clocks using factor analysis can be used study the associations of exposures with biological age acceleration, which can be generalized to include other epigenetic clock combinations. We also empirically compared the regression coefficients using standardized betas for different clocks and adiposity measures. The adiposity measures we used are common, increasing the possibility of replicating our methods; they are also cost-effective and minimally burdensome to the study participants, making them feasible to implement in large cohort studies. Use of a LMIC-based cohort living through rapidly expanding economy gives insights into the generalisation of our findings to other LMIC settings; studying a young and lean population contributes to the body of research on obesity prevention. Adequate adjustment by reproductive factors in females, facilitated by the richness of our dataset, is also a strength, given that this confounder is usually missed.

## Conclusion

Combining biological age acceleration measured by several epigenetic clocks using factor analysis is a valid approach to estimating underlying biological age acceleration. The flexibility of this method can be leveraged to test other combinations of epigenetic clocks. We found consistent associations of adiposity measured by BMI, WC, and WHtR with Factor, PCFactor, PCPhenoAge, PCHannum, and DunedinPACE, with WC and WHtR also being associated with PC GrimAge and PC Horvath 2. DunedinPACE was more sensitive to adiposity in females than males. Replication of this approach in different populations in needed for generalizability.

## Data Availability

All data used in the analysis can be found on https://github.com/rsvolosh/BioAge_Adiposity/. Socioeconomic variables used to calculate household index, sample questionnaires, and all relevant data except for epigenetic clocks (which are available at https://github.com/rsvolosh/BioAge_Adiposity/) can also be found at https://dataverse.unc.edu/dataverse/cebu. Code needed to transform the household asset data into the household wealth index used in this study can be found in the dataverse can be found at https://github.com/jvargh7/cohorts-wealth-gains.

## Acknowledgements

We are grateful to the CLHNS participants whose continued cooperation made this study possible, and to the staff of University of San Carlos Office of Population Studies Foundation Inc. who put forth tremendous efforts to collect, verify, and clean the data used in this study.

## Supporting Information

**S1 Table. Household Assets.**

**S2 Table: Measures of Sampling Adequacy.** Results from KMO tests run clocks used to create Factor (non-PC) and PCFactor (PC).

**S3 Table: Model Fit Statistics for Factor Analysis.**

**S1 Fig: Scree plots.** A: Scree plot for Horvath, Hannum, PhenoAge, GrimAge, DNAmTL, and DunedinPACE. B: Scree plot for PCHorvath2, PCHannum, PCPhenoAge, PCGrimAge, PCDNAmTL, and DunedinPACE.

**S4 Table: Comparison of adiposity measures in parous and nulliparous females.**

**S1 File: Goodness of fit measures for models 1, 2, and final models.**

**S5 Table: Comparison of standardized regression coefficients for Factor, PCFactor, with their component clocks.** For each exposure, regression coefficients for Factor and its components, PCFactor andFactor, and PCFactor and its components were compared using methods outlined in Clogg, Petkova, and Haritou(70).

**S6 Table: Comparison of standardized regression coefficients differing by exposure.** For each outcome associated with more than one measure of adiposity at a statistically significant level, regression coefficients for different measures of adiposity were compared using methods outlined in Clogg, Petkova, and Haritou(70).

